# How do the general population behave with facemasks to prevent COVID-19 in the community?

**DOI:** 10.1101/2020.09.18.20195669

**Authors:** Colin Deschanvres, Thomas Haudebourg, Nathan Peiffer-Smadja, Karine Blanckaert, David Boutoille, Jean-Christophe Lucet, Gabriel Birgand

## Abstract

**IMPORTANCE:** The appropriate use of facemasks, recommended or mandated by authorities, is critical to protect the community and prevent the spread of COVID-19.

**OBJECTIVE:** To evaluate the frequency and quality of facemask use in general populations of different socio-spatial backgrounds.

**DESIGN:** A multi-site observational study carried out from 25 June 2020 to 21 July 2020.

**SETTING:** The observations were carried out in 43 different locations in a region in the west of France, representing various areas: rural and urban, indoor and outdoor, and in areas where masks were mandated or not. An observer was positioned at a predetermined place, facing a landmark, and collected information about the use of facemasks and socio-demographic data.

**PARTICIPANTS:** All individual passing between the observer and the landmark were included.

**EXPOSURE:** The observer collected information on whether a mask was worn, the type of mask used, the quality of the positioning, gender, and the age category of each individual.

**MAIN OUTCOMES AND MEASURES:** The main outcomes were the use of a facemask and the quality of the positioning. Factors associated with these outcomes were identified.

**RESULTS:** A total of 3354 observations were recorded. A facemask was worn by 56.4% (*n=*1892) of individuals, varying from 49% (*n=*1359) in non-mandatory areas and 91.7% (*n=*533) in mandatory areas, including surgical facemasks (56.8%, *n=*1075) and cloth masks (43.2%, *n=*817). The facemask was correctly positioned in 75.2% (*n=*1422) of cases. The factors independently associated with wearing a facemask were being indoors (adjusted odds ratio [aOR], 0.37; 95% confidence interval [CI], 0.31-0.44), being in a mandatory area (aOR, 0.14; 95%CI, 0.10-0.20), female gender (aOR, 0.57; 95%CI, 0.49-0.66), and age >40 years (aOR, 0.54; 95%CI, 0.46-0.63). The factors independently associated with correct mask position were rural location (aOR, 0.76; 95%CI, 0.97-0.98), being in an indoor area (aOR, 0.49; 95%CI, 0.38-0.65), use of a cloth mask (aOR, 0.65; 95%CI, 0.52-0.81), and age >40 years (aOR, 0.61; 95%CI 0.49-0.76).

**CONCLUSIONS AND RELEVANCE:** Information campaigns should promote the use of cloth masks. Young people in general and men in particular are the priority targets. Simplifying the rules to require universal mandatory masking seems to be the best approach for health authorities.

**Key Points:** *Question:* What is the frequency and quality, and their associated factors, of the use of facemasks in general populations of different socio-spatial backgrounds?

*Findings:* Among 3354 observations, 56.4% of individuals wore a facemask, either a surgical mask (56.8%) or a cloth mask (43.2%), and the mask was correctly positioned in 75.2% of cases. Correct use of facemasks was more common in rural and indoors areas, individuals wearing cloth masks, and among those aged >40 years.

*Meaning:* Health authorities should promote the use of cloth masks, engage young people in this process, and consider the mandatory universal masking.

## INTRODUCTION

Since the emergence of the Coronavirus (COVID-19) epidemic, wearing a facemask in the community has become commonplace. In many countries, facemasks are mandatory in crowded areas where social distancing cannot be respected and are recommended outdoors.^1^ Appropriate use of facemasks is critical for protection in the community to prevent the spread of COVID-19.^2^ However, the constraints and discomfort caused in a population unfamiliar with this protective equipment can result in suboptimal use, leading to ineffective protection against COVID-19. Observation and quantification of the quality of facemask use is required to: assess the level of respiratory protection, inform decision makers on the effectiveness of measures, and identify levers for behavior change. We evaluated the frequency and quality of facemask use in general populations with different socio-spatial backgrounds, and contextual factors associated with the appropriate use of the facemask.

## METHODS

From June 25, 2020, to July 21, 2020, we conducted observations in 13 cities and 43 different locations in the Pays de la Loire region in western France with a population of 3.8 million (**Supplementary Figure 1**). The observations were performed in various areas: rural and urban (cities with >10,000 and with <10,000 inhabitants), indoors (shopping centers, train stations) or outdoors (shopping streets), and in areas where masks were or were not mandatory. The observer was positioned in a predetermined place, facing a landmark, and all people passing between the observer and the landmark were included. For each individual, the researcher recorded if a facemask was worn, the type of facemask, and the quality of facemask positioning. The facemask was considered incorrectly worn if it was in one of the positions defined in Table 1. For each observation session, information on the time, location, and mandatory status was recorded. In addition, demographic characteristics were collected, including the gender and age category (21-40, 41-65, and >65 years). The data were collected on a smartphone using a Google form. Contingency tables and chi-squared tests were used for categorical variables. Unadjusted relative risks (ORs) were determined and 95% confidence intervals (95% CI) were computed. Multiple logistic regression was performed. Variables associated with p values <0.25 in the bivariate analysis were entered into the model to obtain maximum likelihood estimates. These analyses were performed using R version 3.6.1.

**Table 1.**
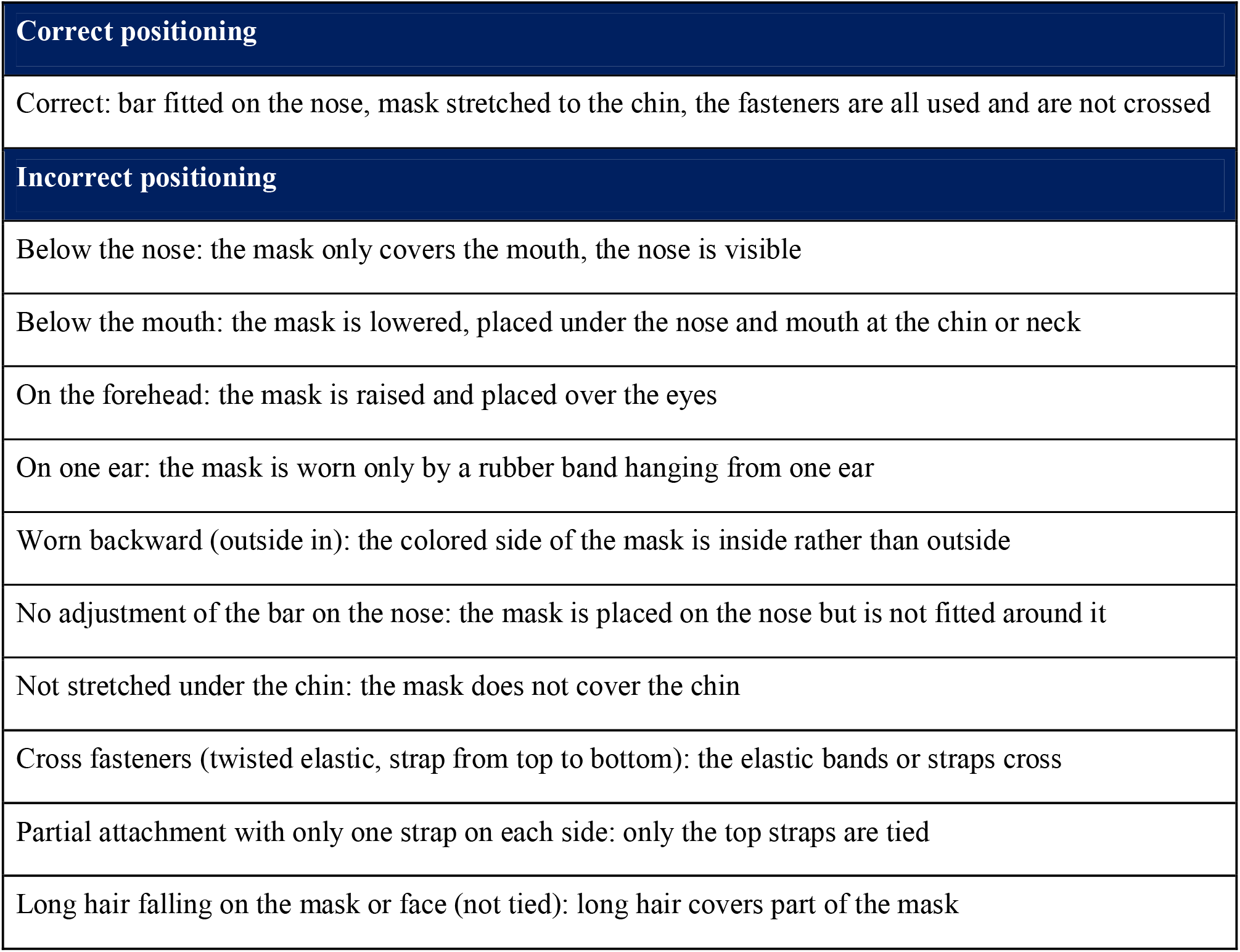
Definitions for the Qualitative Evaluation of Mask Position.

|  |
| --- |
| **Correct positioning** |
| Correct: bar fitted on the nose, mask stretched to the chin, the fasteners are all used and are not crossed |
| **Incorrect positioning** |
| Below the nose: the mask only covers the mouth, the nose is visible |
| Below the mouth: the mask is lowered, placed under the nose and mouth at the chin or neck |
| On the forehead: the mask is raised and placed over the eyes |
| On one ear: the mask is worn only by a rubber band hanging from one ear |
| Worn backward (outside in): the colored side of the mask is inside rather than outside |
| No adjustment of the bar on the nose: the mask is placed on the nose but is not fitted around it |
| Not stretched under the chin: the mask does not cover the chin |
| Cross fasteners (twisted elastic, strap from top to bottom): the elastic bands or straps cross |
| Partial attachment with only one strap on each side: only the top straps are tied |
| Long hair falling on the mask or face (not tied): long hair covers part of the mask |

## RESULTS

A total of 3354 observations were performed during 55 sessions (**Table 2**): 1639 (49%) observations were performed indoors and 1715 (51%) outdoors. The ratio of males to females was 0.73, and 44.6% (*n=*1495) were aged 21-40 years, 35.3% (*n=*1184) were aged 41-65 years, and 20.1% (*n=*675) were >65 years.

**Table 2.** Description of the Study Population, with Demographic Characteristics, Frequency and Qualitative Characteristics of Use of Masks.

| Characteristics | Overall, n (%) | Outdoor, n (%) |  | Indoor, n (%) |  |
| --- | --- | --- | --- | --- | --- |
|  |  | Urban | Rural | Urban | Rural |
| Number of observations | 3354 | 1165 | 550 | 974 | 665 |
| Gender |  |  |  |  |  |
| Female | 1943 (57.9) | 705 (60.5) | 303 (55.1) | 550 (56.5) | 385 (57.9) |
| Male | 1411 (42.1) | 460 (39.5) | 247 (44.9) | 424 (43.5) | 280 (42.1) |
| Age category |  |  |  |  |  |
| 21-40 years | 1495 (44.6) | 705 (60.5) | 141 (25.6) | 456 (46.8) | 193 (29) |
| 41-65 years | 1184 (35.3) | 373 (32) | 190 (34.5) | 365 (37.5) | 256 (38.5) |
| >65 years | 675 (20.1) | 87 (7.5) | 219 (39.8) | 153 (15.7) | 216 (32.5) |
| Time of day |  |  |  |  |  |
| Morning | 1454 (43.4) | 269 (23.1) | 400 (72.7) | 328 (33.7) | 457 (68.7) |
| Afternoon | 1900 (56.6) | 896 (76.9) | 150 (27.3) | 646 (66.3) | 208 (31.3) |
| Mask mandated |  |  |  |  |  |
| No | 2773 (82.7) | 1165 (100) | 550 (100) | 510 (52.4) | 548 (82.4) |
| Yes | 581 (17.3) | 0 (0) | 0 (0) | 464 (47.6) | 117 (17.6) |
| Presence of a facemask |  |  |  |  |  |
| No | 1462 (43.6) | 732 (62.8) | 304 (55.3) | 235 (24.1) | 191 (28.7) |
| Yes | 1892 (56.4) | 433 (37.2) | 246 (44.7) | 739 (75.9) | 474 (71.3) |
| Type of facemask (n = 1892) |  |  |  |  |  |
| Surgical facemask | 1075 (56.8) | 266 (61.4) | 131 (53.3) | 419 (56.7) | 259 (54.6) |
| Cloth mask | 817 (43.2) | 167 (38.6) | 115 (46.7) | 320 (43.3) | 215 (45.4) |
| Quality of mask positioning (n = 1892) |  |  |  |  |  |
| Correct | 1422 (75.2) | 264 (61) | 191 (77.6) | 576 (77.9) | 391 (82.5) |
| Incorrect | 470 (24.8) | 169 (39) | 55 (22.4) | 163 (22.1) | 83 (17.5) |
| Incorrect positioning (n = 470) |  |  |  |  |  |
| Below the mouth | 141 (30) | 82 (48.5) | 14 (25.5) | 40 (24.5) | 5 (6) |
| Below the nose | 130 (27.7) | 37 (21.9) | 23 (41.8) | 37 (22.7) | 33 (39.8) |
| Cross straps | 61 (13) | 8 (4.7) | 6 (10.9) | 31 (19) | 16 (19.3) |
| Not adjusted on the nose | 43 (9.1) | 10 (5.9) | 3 (5.5) | 24 (14.7) | 6 (7.2) |
| Hair down on face | 33 (7) | 17 (10.1) | 5 (9.1) | 5 (3.1) | 6 (7.2) |
| Partial mask attachment with strap | 35 (7.4) | 9 (5.3) | 1 (1.8) | 13 (8) | 12 (14.5) |
| Not stretched under the chin | 13 (2.8) | 2 (1.2) | 1 (1.8) | 8 (4.9) | 2 (2.4) |
| On one ear | 10 (2.1) | 1 (0.6) | 2 (3.6) | 4 (2.5) | 3 (3.6) |
| On the forehead | 3 (0.6) | 3 (1.8) | 0 (0) | 0 (0) | 0 (0) |
| Worn backward | 1 (0.2) | 0 (0) | 0 (0) | 1 (0.6) | 0 (0) |

A facemask was worn by 56.4% (*n=*1892) of individuals, varying from 40% (*n=*679) outdoors and 74% (*n=*1213) indoors, 59% (*n=*720) in rural areas, 55% (*n=*1172) in urban areas, 49% (*n=*1359) in non-mandatory areas, and 91.7% (*n=*533) in mandatory areas. With regard to the type of facemask worn, 56.8% (*n=*1075) wore a surgical facemask and 43.2% (*n=*817) wore a cloth mask. Among the 1892 individuals wearing a facemask, 75.2% (*n=*1422) were wearing it correctly. Of the 470 masks positioned incorrectly, 141 (30%) were below the chin and 130 (27.7%) below the nose. Overall, 42.4% (*n=*1422 of 3354) of the population studied was effectively protected.

In the multivariate analysis, facemasks were significantly more often worn indoors (adjusted odds ratio [aOR], 0.37; 95%CI, 0.31-0.44; p<0.001) and in mandatory areas (aOR, 0.14; 95%CI, 0.10-0.20; p<0.001). Facemasks were significantly less frequently worn by males (aOR, 1.75; 95%CI, 1.51-2.04; p<0.001) and by younger individuals aged 21-40 years (aOR, 2.28; 95%CI, 1.83-2.85; p<0.001) and those aged 41-65 years (aOR, 1.34; 95%CI, 1.08-1.68; p=0.008) (**Table 2**).

Among the individuals wearing a facemask, correct positioning was significantly higher in rural (aOR, 0.75; 95%CI, 0.57-0.97; p=0.03) and indoor areas (aOR, 0.49; 95%CI, 0.38-0.65; p<0.001). The use of cloth masks in comparison with surgical masks was significantly associated with correct positioning (aOR, 0.65; 95%CI, 0.52-0.81; p<0.001). Incorrect positioning was significantly associated with the younger age group (21-40 years) (OR, 1.47; 95%CI, 1.10-1.98; p=0.01) (**Table 3**).

**Table 3.** Univariate and Multivariable Analysis of Factors Influencing the Use and the Visual Correct Position of Facemask Fit.

| Factors | Facemask | No facemask | Univariate OR (95% CI) | p | Multivariate aOR (95% CI) | p | Correct position | Incorrect position | Univariate OR (95% CI) | p | Multivariate aOR (95% CI) | p |
| --- | --- | --- | --- | --- | --- | --- | --- | --- | --- | --- | --- | --- |
| **Number** | 1892 (56.4) | 1462 (43.6) |  |  |  |  | 1422 (75.2) | 470 (24.8) |  |  |  |  |
| **Area** |  |  |  |  |  |  |  |  |  |  |  |  |
| Urban | 1172 (54.8) | 967 (45.2) | Reference |  | Reference |  | 840 (71.7) | 332 (28.3) | Reference |  | Reference |  |
| Rural | 720 (59.3) | 495 (40.7) | 0.83 (0.72-0.96) | 0.012 | 0.85 (0.71-1.02) | 0.075 | 582 (80.8) | 138 (19.2) | 0.6 (0.48-0.75) | <0.001 | 0.75 (0.57-0.97) | 0.03 |
| **Location** |  |  |  |  |  |  |  |  |  |  |  |  |
| Outdoor | 679 (39.6) | 1036 (60.4) | Reference |  | Reference |  | 455 (67) | 224 (33) | Reference |  | Reference |  |
| Indoor | 1213 (74) | 426 (26) | 0.23 (0.2-0.27) | <0.001 | 0.37 (0.31-0.44) | <0.001 | 967 (79.7) | 246 (20.3) | 0.52 (0.42-0.64) | <0.001 | 0.49 (0.38-0.65) | <0.001 |
| **Mandatory** |  |  |  |  |  |  |  |  |  |  |  |  |
| No | 1359 (49) | 1414 (51) | Reference |  | Reference |  | 1016 (74.8) | 343 (25.2) | Reference |  | Reference |  |
| Yes | 533 (91.7) | 48 (8.3) | 0.09 (0.06-0.12) | <0.001 | 0.14 (0.1-0.2) | <0.001 | 406 (76.2) | 127 (23.8) | 0.93 (0.73-1.17) | 0.52 | 1.2 (0.89-1.62) | 0.23 |
| **Time of day** |  |  |  |  |  |  |  |  |  |  |  |  |
| Morning | 800 (55) | 654 (45) | Reference |  | Reference |  | 641 (80.1) | 159 (19.9) | Reference |  | Reference |  |
| Afternoon | 1092 (57.5) | 808 (42.5) | 0.91 (0.79-1.04) | 0.16 | 0.79 (0.66-0.94) | 0.007 | 781 (71.5) | 311 (28.5) | 1.61 (1.29-2) | <0.001 | 1.18 (0.92-1.52) | 0.20 |
| **Type of mask** |  |  |  |  |  |  |  |  |  |  |  |  |
| Surgical |  |  |  |  |  |  | 770 (71.6) | 305 (28.4) | Reference |  | Reference |  |
| Cloth |  |  |  |  |  |  | 652 (79.8) | 165 (20.2) | 0.64 (0.51-0.79) | <0.001 | 0.65 (0.52-0.81) | <0.001 |
| **Gender** |  |  |  |  |  |  |  |  |  |  |  |  |
| Female | 1190 (61.2) | 753 (38.8) | Reference |  | Reference |  | 896 (75.3) | 294 (24.7) | Reference |  | Reference |  |
| Male | 702 (49.8) | 709 (50.2) | 1.6 (1.39-1.83) | <0.001 | 1.75 (1.51-2.04) | <0.001 | 526 (74.9) | 176 (25.1) | 0.98 (0.79-1.22) | 0.86 | 1.03 (0.82-1.29) | 0.79 |
| **Age category** |  |  |  |  |  |  |  |  |  |  |  |  |
| >65 years | 451 (66.8) | 224 (33.2) | Reference |  | Reference |  | 357 (79.2) | 94 (20.8) | Reference |  | Reference |  |
| 41-65 years | 724 (61.1) | 460 (38.9) | 1.28 (1.05-1.56) | 0.015 | 1.34 (1.08-1.68) | 0.008 | 578 (79.8) | 146 (20.2) | 0.96 (0.72-1.29) | 0.78 | 0.85 (0.63-1.15) | 0.28 |
| 21-40 years | 717 (48) | 778 (52) | 2.18 (1.81-2.64) | <0.001 | 2.28 (1.83-2.85) | <0.001 | 487 (67.9) | 230 (32.1) | 1.79 (1.36-2.37) | <0.001 | 1.47 (1.1-1.98) | 0.01 |

Outcome was assessed using multivariate logistic regression estimating odd ratios (ORs) and 95% confidence intervals (CIs). Crude and adjusted odds ratios determine the factors associated with the use and the visual correct positioning of facemask fit. Values presented as number (%). $p < 0.05$ was considered statistically significant. The multivariate model had no missing data.

## DISCUSSION

In a post lockdown context with large clusters of COVID-19 cases leading to a potential second wave, less than half of the individuals were correctly protected in the general population. Unsurprisingly, the mandatory process was the most powerful variable associated with increased use of facemasks. The mandatory approach may represent the best political lever to increase the level of facemask use in the general population.

Among the people wearing a facemask incorrectly, the most commonly observed positions were below the chin or below the nose. Following the mandate to wear a facemask in shops and indoor public areas, people going from one shop to another are not keeping their facemask on. These observations suggest that facemasks are being handled and repositioned by individuals going between two mandatory areas, perhaps due to respiratory discomfort. These behaviors could lead to an increase in the risk of transmission, particularly through hand contamination. This fact is important due to the difficulty in complying with hand hygiene measures when putting the facemask on and taking it off. Mandatory universal masking, even in the absence of scientific evidence outdoors, has the advantage of simplifying the measure and avoiding misuse.

The positioning of cloth masks was significantly better in comparison with surgical facemasks. People wearing cloth masks may correspond to a sub-population of individuals engaged by choosing their mask, the cloth, and pattern. On the other hand, the characteristics of surgical facemask (impersonal, single use, more expensive, potentially less comfortable to wear) decrease compliance with best practices. Good quality cloth masks that can be customized with various sizes and patterns may adapt better to the face, making them more comfortable, and may lead to better engagement by users.^3^

The use of facemasks was significantly lower and more often worn incorrectly in the population <40 years and in males independently of non-use of the mask. This finding is consistent with the increase in COVID-19 cases in the younger population during the post lockdown period.^4,5^ The lack of clinical impact associated with the social factors that applied during the lockdown potentially led to lower adherence to preventive measures by the young population during the summer break. These populations represent a target for authorities in their information campaigns to optimize the protection of the general population. Facemasks were worn correctly by those in rural areas compared with urban areas. In small cities, people are living together as part of an identifiable network, with significant social norms and better individual behaviors. In contrast, in urban populations, individuals are anonymous, with less reference to norms and altruistic measures.

To our knowledge, this study is the first to quantify the frequency and quality of the use of facemasks in the general population. However, the visual and potentially subjective evaluation of the criteria should be acknowledging as potential limitations. Despite the inclusion of a range of situations at the regional scale, the generalizability is questionable.

## CONCLUSION

Information campaigns should promote the use of cloth masks. Young people in general and men in particular are the priority targets. Simplifying the rules to require universal mandatory masking seems to be the best approach for health authorities.

## Data Availability

All data will be available to those who make the request.

## Declarations

### Ethics Approval and Consent to Participate

Not applicable.

### Consent for Publication

Not applicable.

### Availability of Data and Material

Data sharing not applicable to this article because no datasets were generated or analyzed during the study.

## Acknowledgments

None.

